# Association and Timing of Changes in Emotion Regulation and Mood along the Menstrual Cycle in Women with Premenstrual Syndrome

**DOI:** 10.1101/2025.03.18.25324201

**Authors:** Belinda Pletzer, Anna Gnaiger, Julia Kinzelmann, Patricia Werlein, Livia Rauter, Esmeralda Hidalgo-Lopez

**Affiliations:** Department of Psychology, University of Salzburg, Salzburg, AUSTRIA; Centre for Cognitive Neuroscience, University of Salzburg, Salzburg, Austria; Department of Psychology, University of Michigan, Ann Arbor, Michigan, USA; Chronic Pain and Fatigue Research Center, University of Michigan Medical School, Ann Arbor, Michigan, USA

**Author notes:** corresponding author: DDr. Belinda Pletzer Centre for Cognitive Neuroscience University of Salzburg Hellbrunnerstr. 34, 5020 Salzburg, AUSTRIA.

## Abstract

Premenstrual dysphoric disorder (PMDD) is characterized by the recurrence of major psychiatric symptoms in the final week before the onset of menses, which severely impact women’s quality of life. The etiology of PMDD, or more generally premenstrual syndrome (PMS) including subclinical forms, is not well understood and both biological and psychological models have been postulated. In the current manuscript we explore the role of emotion regulation ability for the emergence of premenstrual symptoms. Specifically, we were interested whether difficulties in emotion regulation in women with PMS/PMDD persisted as a trait across all cycle phases or were specific to the symptomatic premenstrual phase. We also explored for the first time, whether difficulties in emotion regulation already emerged during the high progesterone mid-luteal phase. 105 women aged 18 to 35 years completed a trait questionnaire on difficulties in emotion regulation, as well as two state measures of emotion regulation during three cycle phases (mid-follicular, mid-luteal, premenstrual). PMS/PMDD was prospectively confirmed across two menstrual cycles in 54 women, while the remaining 51 were assigned to the control group. Our results confirm previous findings that women with PMS/PMDD subjectively report more difficulties in emotion regulation on a trait scale. However, these difficulties were not confirmed in any of the state measures employed in the current study. Furthermore, difficulties in emotion regulation did not account for premenstrual symptom severity in a mediation analysis. This raises the question, whether difficulties in emotion regulation underly premenstrual symptoms or the perception of impaired emotion regulation arises as a consequence of untreated PMS/PMDD. Noteworthy, we were also able to demonstrate via Bayesian analyses, that mood worsening in the premenstrual phase is not a universal occurrence and absent in healthy controls. Taken together these findings highlight the importance of adequate diagnosis and treatment of PMDD.

## Introduction

Premenstrual dysphoric disorder (PMDD) is characterized by the repeated occurrence of at least one out of four major psychiatric symptoms in the final week before the onset of menses, which subsequently subside in the three days after onset of menses. The key symptoms include depression, anxiety, irritability and mood lability. Similar to other affective disorders, these symptoms are accompanied by sleep disturbances, fatigue, difficulty concentrating and a general loss of interest in everyday activities (Halbreich et al., 2003). A total of five symptoms are required to recur in two consecutive cycles to meet the criteria for PMDD according to the fifth edition of the Diagnostic and Statistical Manual of Mental Disorders (DSM-V). The prevalence of PMDD is about 3-8% (Halbreich et al., 2003; Zhu et al., 2024). However, serious impairments in quality of life can also occur with fewer symptoms. Therefore, subclinical symptoms restricted to the premenstrual phase are often summarized and studied as premenstrual syndrome (PMS). Due to the lack of diagnostic cut-off criteria, as well as potential cultural differences, prevalence of PMS varies across studies and countries, ranging from 9-57% (Zhu et al., 2003). In Western countries, PMS and PMDD are rarely diagnosed (Weisz et al., 2009), even though symptoms significantly impact the affected women’s quality of life and increase the risk of suicide (Prasad et al., 2021; Zhu et al., 2024). Given the potential severe personal, but also socio-economical consequences, understanding the psychobiological factors contributing to the etiology of premenstrual symptoms is of utmost importance.

Premenstrual symptoms appear to be triggered by elevated progesterone levels in the mid-luteal cycle phase, as evidenced by the fact that premenstrual symptoms disappear when progesterone fluctuations are abolished via GnRH agonists and re-appear when progesterone is re-substituted (Sundstrom-Poromaa et al., 2020). Nevertheless, a direct relationship between symptom severity and progesterone concentration is not evident, which is further reinforced by the fact that hormonal profiles along the menstrual cycle do not appear to differ between women with and without PMDD (Bäckström et al., 2003). Rather, premenstrual symptoms seem to characterize differential reactivity to typical progesterone elevations in the mid-luteal cycle phase. Various avenues regarding the underlying causes of this altered response have been explored including biological models, such as genetic polymorphisms related to hormone sensitivity (Huo et al., 2007; Lahnsteiner et al., 2025), or altered neurophysiology, particularly in the dopaminergic and serotonergic system (Nappi et al., 2022). Psychological models on the other hand postulate alterations in emotion regulation capacity or emotion regulation strategies as the underlying cause of altered emotional responses to hormonal fluctuations along the menstrual cycle in women with premenstrual syndrome.

These models are of course not mutually exclusive. Emotion regulation summarizes all cognitive processes designed to influence the intensity or duration of emotional experiences with the goal to adapt to environmental challenges (Aldao and Plate, 2018). As such, emotion regulation may reflect brain connectivity patterns affected by altered neurophysiology. In fact, the luteal phase of the menstrual cycle has been termed a window-of vulnerability for the emergence of stress-related disorders in women based on the activation and connectivity patterns of three distinct brain networks (Andreano et al., 2018). In the luteal phase of the menstrual cycle, increased salience for negative emotional stimuli has been reported both at the behavioral (Lusk et al., 2017; Guevarra et al., 2023) and neuronal level, reflected in increased activation and connectivity within the salience network (Engman et al., 2018; Hidalgo-Lopez et al., 2021). At the same time an increased tendency towards self-referential processing has been postulated by findings at the neuronal level, reflected by increased connectivity within the default mode network (Petersen et al., 2014; Hidalgo-Lopez et al., 2021). A heightened attribution of negative emotional experiences to oneself is at the core of several psychiatric symptoms, such as depression (see Collins et al., 2023 for a meta-analysis). We recently observed, that in healthy women this increased connectivity of the salience and default mode network during the luteal cycle phase is generally balanced by altered top-down control over those two networks via the executive control network (Hidalgo-Lopez et al., 2021). It has been demonstrated that effective emotion regulation relies on the interplay between those networks, with a crucial role for the executive control network, particularly prefrontal areas (Menon et al., 2019; Etkin et al., 2015).

However, while neuroimaging evidence regarding connectivity shifts during the mid-luteal phase is converging, only few studies have studied emotion regulation along the menstrual cycle in healthy women at a behavioral level (Wu et al., 2014; Olatunje et al., 2020; Doornweerd et al., 2024). Noteworthy, these studies have focused on reappraisal, an emotion regulation strategy that reframes a situation to alter its emotional impact (Ford et al., 2017) and depends on the effective interplay between the salience and executive control networks (Etkin et al., 2015; Cutuli, 2014). Reappraisal is often considered an adaptive emotion regulation strategy and has been studied in opposition to suppression, which involves the inhibition of emotional expression rather than the regulation of the emotional experience itself (Cutuli, 2014). Wu et al. (2014) and Doornweerd et al. (2024) found no menstrual cycle changes in an objective emotion regulation task, but a reduction in subjective reappraisal success ratings during the mid-luteal cycle phase. Conversely, Olatunje et al. (2020) observed no menstrual cycle related changes in subjective reappraisal and suppression but found suppression to be more effective than reappraisal in a disgust regulation task during the follicular phase. Given these inconsistencies across studies, it remains unclear whether the effectiveness of emotion regulation or the preference for certain emotion regulation strategies changes along the menstrual cycle. It is thus unclear whether menstrual cycle-related changes in connectivity between large-scale brain networks represent a healthy adaptation to hormonal fluctuations and serve to preserve the effectiveness of emotion regulation or whether they underlie changes in the effectiveness of emotion regulation. Likewise, it can be debated whether these connectivity changes prevent premenstrual mood symptoms in healthy women and are thus absent in women with PMS/PMDD or vice versa, whether these alterations are present in all women and contribute to mood symptoms in women with PMS/PMDD.

Unlike the menstrual cycle per se, PMS and PMDD have been frequently studied regarding their association to emotion regulation. Most studies addressed the question from a trait perspective, i.e. via a single assessment using a trait questionnaire without controlling for menstrual cycle phase. These studies consistently demonstrate more difficulties in all domains of emotion regulation and reduced use of reappraisal in women with PMS/PMDD compared to healthy controls (Wu et al., 2016; Manikandan et al., 2016; Petersen et al., 2016; Nayman et al., 2022; Nasiri et al., 2022; Lin et al., 2022; Elazar et al., 2023). What remains unclear is whether altered emotion regulation in women with PMS/PMDD represents a state emerging in the late luteal cycle phase or is in fact a trait that allows mood symptoms to emerge during the late luteal cycle phase. Answering this question at the behavioral level may shed some light on the debate surrounding the functionality of connectivity changes along the menstrual cycle and may thus aid the optimal design of future neuroimaging experiments. If altered inter-network connectivity during the mid-luteal cycle phase represents a healthy adaptation to hormonal fluctuations, we expect them to be absent in women with PMS/PMDD, resulting in constant emotion regulation impairments along the menstrual cycle. However, if altered inter-network connectivity underlies the emergence of mood symptoms in the late luteal cycle phase, we expect emotion regulation impairments only in the late luteal cycle phase.

The few studies approaching the question from a state perspective have included the asymptomatic mid-follicular phase, as well as the symptomatic late-luteal phase, but have arrived at vastly different results. Questionnaire approaches found more problems in emotion regulation in women with PMS/PMDD, irrespective of cycle phase (Eggert et al., 2016; Yen et al., 2018). However, it is possible that explicit self-report questionnaires do not capture state variations in emotion regulation, given that women have experienced those difficulties for a long period of time and may thus attribute them as an integral part of their personality, even if the problems emerge only cyclically. To address this question, a task requiring the active implementation of emotion regulation strategies under laboratory conditions might provide more objective measures of emotion regulation capacity. Using an affect misattribution procedure, Eggert et al. (2016) obtained only very weak indication of implicit emotion regulation ability being selectively impaired in the late luteal cycle phase. However, affect misattribution measures the misattribution of emotional reactivity rather than emotion regulation per se. Using an explicit emotion regulation task, Petersen et al. (2018) observed emotion regulation ability to be selectively impaired during the late luteal phase in women with PMS/PMDD along with hypo-activation in the dorsolateral prefrontal cortex during an emotion regulation task (Petersen et al., 2018). Interestingly, no study to date has explored emotion regulation in women with PMS/PMDD during the mid-luteal cycle phase, even though progesterone appears to be the trigger for premenstrual symptoms and connectivity shifts alike. It is thus possible that emotion regulation impairments that occur in the mid-luteal cycle phase contribute to the later emergence of premenstrual symptoms.

To address this question and identify whether in women with PMS/PMDD emotion regulation is consistently impaired across the menstrual cycle or selectively in either the mid-or late-luteal cycle phase, we studied emotion regulation during three different cycle phases (mid-follicular, mid-luteal and late-luteal) in women with PMS/PMDD and healthy controls. Given that the emotion regulation task employed by Petersen et al. (2018) proofed to be most sensitive to intra-individual variations in previous investigations (Petersen et al., 2018; Silvers et al., 2012), we chose to implement this task combined with emotion regulation questionnaires used in previous studies (Wu et al., 2016; Petersen et al., 2016; Nasiri et al., 2021). We also explore, whether emotion regulation impairments or other factors, like physical symptoms or perceived stress, mediate premenstrual symptom severity in the late luteal cycle phase of women with PMS/PMDD.

## Methods

### Participants

Power analyses suggest a required number of 86 participants for this study to detect group differences of moderate effect size (f = 0.25) with 80% power among three repeated measurements. Accounting for 25% dropouts, we aimed to include 108 participants, 54 with suspected PMS/PMDD and 54 controls. 129 participants were recruited and after attrition (see procedure), 105 participants were analyzed.

Participants were included in the study if they were female, nulliparous, aged between 18 and 35 years, were not on hormonal contraceptives and demonstrated a regular menstrual cycle with an average duration of 21 to 35 days and no more than 7 days variability between individual cycle lengths (compare Fehring et al., 2006) based on self-reports of their past three cycle dates. Participants were excluded from the study, if they reported endocrinological or neurological disorders, medication use or hormonal contraceptive use in the past six months, or hospitalization for psychological disorders in the past six months.

### Ethics statement

The study was approved by the University of Salzburg’s ethics committee (GZ 24/2022). All participants provided written informed consent to partake in the study. All methods conform to the Code of Ethics of the World Medical Association (Declaration of Helsinki).

### Procedure

In an initial screening session, participants signed informed consent, provided demographic information, and completed the PSST as well as trait questionnaires on mood and emotion regulation. The expected onset of next menses was calculated based on previous cycle dates and three test sessions were scheduled in three different cycle phases, order counterbalanced. One session was scheduled in the mid-follicular phase, i.e. cycle days 6 to 10, one session in the mid-luteal phase, i.e. 11 to 4 days before the expected onset of next menses and one session in the late luteal phase, i.e. within the last 3 days before the expected onset of next menses. Cycle phases were confirmed by next period onset, and scheduling of test sessions was adapted throughout the study to adjust for deviations between actual and expected cycle length. For example, if the first session was expected to be scheduled in the late luteal phase, but the onset of next menses was later than expected, such that the session was in fact scheduled in the mid-luteal phase, the dates of the next test sessions were adjusted accordingly to capture the remaining two cycle phases.

Of the 129 participants recruited for this study, 10 dropped out after the first test session. 14 participants were excluded because of long and/or anovulatory cycles during the study (n = 8) or incomplete and/or inconsistent DRSP information (n = 6). Accordingly, data from 105 participants were included in the analyses. Among 65 participants test sessions from all three cycle phases were included. Of the remaining 40 participants, scheduled sessions for either the late luteal (n = 22, i.e., 21%) or the mid-luteal (n = 18, i.e., 17%) phase missed the critical window and were thus excluded from analyses (n = 22) or recoded (n = 18). Thus, four participants entered analyses with two late luteal test sessions, while 14 participants entered analyses with two mid-luteal test sessions. In total, 104 mid-follicular (average cycle day = 7.38 ± 2.15), 100 mid-luteal (average cycle day = 6.68 ± 1.85) and 88 late luteal sessions (average cycle day = -1.80 ± 0.86) were included in the analyses. After exclusions, 37 participants had their first test session in the mid-follicular phase, 28 in the mid-luteal phase and 35 in the late luteal phase.

During each test session participants completed two tasks on executive functions, which are described elsewhere (Gnaiger et al., in prep.), followed by (i) the emotion regulation task, (ii) the emotion regulation questionnaire and (iii) state mood questionnaires. Furthermore participants provided a total of five saliva samples per session for hormone analysis, scheduled at the beginning, the end and in between each experimental block.

### Assessment of premenstrual symptoms

To balance recruitment, premenstrual symptom severity was initially screened using the premenstrual symptom screening tool (PSST), however final group allocation was based on daily ratings of severity of problems (DRSP) ratings over two consecutive cycles.

#### Premenstrual symptom screening tool (PSST)

The *Premenstrual Symptom Screening Tool* (PSST, Steiner et al., 2003) assesses how strongly participants experience symptoms like depression, anxiety, mood lability, irritability, problems concentrating, sleep problems, fatigue, loss of interest as well as physical symptoms during the premenstrual phase (14 items) and how strongly they perceive these symptoms impact their daily life (5 items) on a 4-point Likert scale (1 = not at all, …, 4 = severe). In addition, the questionnaire assesses whether participants had ever received a diagnosis of PMS/PMDD by a clinician. A PMDD diagnosis is suspected when at least one of the core psychological symptoms (depression, anxiety, mood lability, irritability) is rated as severe, a total of five symptoms are rated as moderate to severe, and at least one impact item is rated as severe. If a total of five symptoms are rated as moderate to severe, but core psychological symptoms and impact ratings are only moderate, a milder form of PMS is suspected.

#### Daily rating of severity of problems (DRSP)

The *Daily Rating of Severity of Problems* (DRSP, Endicott et al., 2006) is a daily scoring tool for participants to track common premenstrual symptoms and their impact on daily life on a 6-point Likert scale on a daily basis. The symptom detail can be adjusted with the minimum number of items representing the 11 PMDD symptoms according to the DSM-V. In the current study, we included the 11 items representing DSM-V symptoms, as well as 5 additional items for more detailed assessment of physical symptoms, resulting in a total of 16 items. Summary scores were computed for the 11 items assessing psychological symptoms, as well as the five items assessing physical symptoms. Symptom severity was assessed by calculating the percent increase from cycle days 6 to 10 (mid-follicular phase) to the five premenstrual days over two consecutive cycles for each individual symptom. Participants were assigned to the PMDD group if at least five symptoms, one being a core psychological symptom (depression, anxiety, mood swings, irritability), increased by more than 50% in the premenstrual phase during both cycles. Participants were assigned to the PMS group if at least three symptoms increased by more than 50% and at least one core psychological symptom increased by 30% in the premenstrual phase during both cycles. Otherwise, participants were assigned to the control group.

### Trait depression and anxiety

To differentiate cyclically modulated mood symptoms from trait depression and anxiety, depression scores were assessed using the German version of the *Becks Depression Inventory* (BDI-II, Hautzinger et al., 2006), whereas trait anxiety was assessed using the German version of the *Becks Anxiety Inventory* (BAI, Margraf & Ehlers, 2007). These instruments were designed to assess the severity of depression and anxiety disorders covering the respective diagnostic criteria according to the DSM-V (APA, 2013). Both contain 21 items to be rated on a 4-point Likert scale. While the BDI-II provides participants with a choice between four statements expressing varying symptom strength (0-3), in the BAI psychological and somatic anxiety symptoms are rated according to their severity from not at all (0) to severe (3). Sum scores are calculated over the 21 item responses, thus the maximum score possible is 63 on both scales.

### State Mood Questionnaires

During each test session, participants completed the *Positive and Negative Affect Schedule* (PANAS, Watson, 1988; German version: Krohne et al., 1996), state version of the *State Trait Anxiety Inventory* (STAI, Spielberger et al., 1971; German version: Laux et al., 1981), as well as the Perceived Stress Scale (PSS, Schneider et al., 2020).

The PANAS is a well-validated 20-item instrument consisting of 10 positive and 10 negative affective adjectives, for which participants rate how much they apply to their current mood on a 5-point Likert scale ranging from 1 “not at all” to 5 “very much”. In the current sample, Cronbach’s alpha was 0.89 for positive affect and 0.84 in negative affect.

The state scale of the STAI is a well-validated 20-item instrument including adjectives related to the psychological or physical symptoms of anxiety or nervousness or their opposite, i.e., relaxation. Participants rate on a 4-point Likert scale, how much each adjective describes their current emotional state. In the current sample Cronbach’s alpha for the STAI was 0.90.

The PSS consists of 10 items addressing how often participants felt helpless when faced with stressful situations. For the current study, the items were adapted to ask about participants’ thoughts and feelings in the last days rather than the last month. Participants rate each item on a 5-point Likert scale from 1 “not at all” to 5 “very often”. Subscales for helplessness and self-efficacy can be calculated. In the current sample, Cronbach’s alpha was 0.87 for the overall PSS score, 0.82 for helplessness and 0.75 for self-efficacy.

### Emotion Regulation Assessment

In order to obtain a comprehensive picture of participants’ emotion regulation abilities, we chose to combine (i) an extensive trait-instrument assessing various aspects of emotion regulation strategies in detail, (ii) a short state instrument assessing two selective emotion regulation strategies, as well as (iii) a task assessing the situational success of emotion regulation irrespective of the strategy used.

#### Difficulties in emotion regulation scale (DERS)

The DERS (Gratz & Roemer, 2004; German Version: Gutzweiler & In-Albon, 2019) is a 36-item instrument designed to assess difficulties in four domains of emotion regulation, i.e. awareness of emotions, acceptance of emotions, ability for goal-directed behavior and effectiveness of emotion regulation strategies. Participants rate each item on a 5-point Likert scale from 1 “almost never” to 5 “almost always”. Thus, higher scores indicate stronger difficulties in emotion regulation. In addition to the sum score across the 36 items, a factor analysis (Gutzweiler & In-Albon, 2019) suggests that subscale scores can be formed for (i) the non-acceptance of emotional reactions, (ii) difficulties in goal-directed behavior, (iii) impulse control difficulties, (iv) the lack of ability to perceive one’s own emotions, i.e., emotional awareness, (v) the limited access to emotion regulation strategies and (vi) the lack of emotional clarity. In the current sample, Cronbach’s alpha was 0.95 for the total score and ranged from 0.76 to 0.89 for subscale scores.

#### Emotion regulation Questionnaire (ERQ)

The ERQ is one of the first validated instruments for emotion regulation assessment (Gross & John, 2003; German Version, Abler & Kessler, 2009) and was specifically designed to assess the preferential use of one of two emotion regulation strategies, i.e. reappraisal and suppression. It consists of 10 items belonging to two subscales, which are each rated on a 7-point Likert scale from 1 “not at all true” to 7 “completely true”. The subscale for reappraisal is comprised of 6 items assessing how strongly participants try to change their way of thinking or re-evaluate emotions. The subscale for suppression consists of four items assessing how strongly participants try to suppress the expression of emotions (Preece et al., 2020). In the current sample Cronbach’s alpha was 0.83 for the reappraisal subscale and 0.64 for the suppression subscale.

#### Emotion Regulation Task (ERT)

The emotion regulation task was modelled after Silvers et al (2012), though three different task versions were constructed to avoid repetition of emotional images across test sessions. The task was implemented in Presentation (Neurobehavioral Systems, http://www.neurobs.com/). During each test session, participants were presented with 20 neutral and 20 negative emotional images selected from the International Affective Picture System (IAPS), in randomized sequence. Arousal and valence scores as reported in Lang et al. (1997) were matched across test versions. Prior to the presentation of each image participants were instructed via the cues “close” or “far” to either create emotional closeness to the presented image by imagining that they were standing close to the scene depicted and to allow themselves to experience all the emotions that the photo evoked or to emotionally distance themselves from the presented image by imagining they were further away from the scene and focus less on emotional content and more on the facts of the photo. Participants were trained in these proximal and distal perspective taking strategies prior to performing the task. The cues were counterbalanced across negative and neutral images and presented for 2 seconds, followed by the presentation of images for 8 seconds. Following a 3 second interstimulus interval, participants were asked to rate the intensity of their negative emotions on a nine-point scale (1 = very low negative affect to 9 = very high negative affect) by pressing the corresponding number on the keyboard. The task allows for the calculation of an emotional reactivity score by calculating the percent increase in negative affect from the close neutral to the close negative image condition ([close negative – close neutral]/close neutral × 100). Emotion regulation is calculated as percent reduction in negative affect from the close negative to the far negative image condition ([close negative – far negative]/close negative × 100).

### Hormonal assessment

Five saliva samples of approximately 2ml were collected via the passive drool method throughout each session and stored at -20°. To remove solid particles prior to analysis, samples were centrifuged twice for 15 and 10 min respectively at 3000 rounds per minute using and Eppendorf 5702 centrifuge. To control for pulsatile hormone release, the supernatant was pooled across the five samples per session. Salivary estradiol and progesterone were assessed using Salimetrics ELISA kits. Each pooled sample was analysed twice and in case the coefficient of variance in a sample superseded 25%, analysis of all samples of the respective participant was repeated.

### Statistical analysis

Statistical analyses were performed in R 4.2.3 and RStudio 2024.04.2. Trait measures were compared between groups using Mann Whitney U Tests, while state measures were analysed in the context of linear mixed effects models via the *lme* function of the *nlme* package (Pinheiro et al., 2017). Dependent variables were visually checked for normality and outliers prior to analysis. In case of right skewed distributions, dependent variables were log-transformed prior to analyses. Outliers were removed if they exceeded the mean of the respective cycle phase by more than three standard deviations. All models included menstrual cycle phase, group, and their interaction as fixed effects, as well as participant number as random effect. Significance of fixed effects was determined using the *anova* function of the *stats* package followed by pairwise Tukey tests using the *glht* function of the *multcomp* package (Hothorn et al., 2008). In case of significant interactions, separate lme’s evaluating the effect of menstrual cycle phase were performed for the PMS/PMDD and control group. Dependent variables were scaled, such that b-values represent standardized effect sizes based on standard deviations comparable to Cohen’s d. P-values were FDR-corrected for multiple comparisons across all models.

In order to assess the probability of the null hypothesis relative to the alternative hypothesis in case of non-significant effects, Bayes factors in support of the null hypothesis were calculated using the *lmBF* function of the *BayesFactor* package (Morey et al., 2015). Specifically, we calculated the Bayes factor for a model without (i) the interaction of group and cycle phase, (ii) the main effect of cycle phase and (iii) the main effect of group relative to the respective models without those terms (BF_01_) using 10.000 iterations for Monte Carlo sampling. Bayes factors indicate the likelihood of one model relative to another model. For example, a BF_01_ of 3 indicates that the null hypothesis, i.e., the model without a certain term, is three times more likely than the alternative hypothesis, i.e., the model including a certain term. A BF_01_ of 0.33 on the other hand, indicates that the alternative hypothesis is 3 times more likely than the null hypothesis. If Bayes factors indicated a likelihood above 3 for including the interaction between group and menstrual cycle in the model, Bayes factor for the main effects of group and cycle were not evaluated, but Bayes factors for the main effect of cycle were evaluated separately in each group.

In order to explore, whether psychological premenstrual symptoms were attributable to hormone measures, mood or emotion regulation abilities, we performed mediation analyses for the group differences in psychological premenstrual symptoms in the late luteal phase using bootstrapped samples via the *mediation* package.

## Results

### Final sample

105 participants fulfilling all inclusion criteria completed the study. Participants were aged between 19 and 35 years (mean age = 24.35, SD = 3.67), the majority had passed general qualification for university entrance and was still in education. Following scoring rules for the DRSP, 54 participants were assigned to the control group and 51 participants were assigned to the PMS/PMDD group. Among the latter, 29 fulfilled the criteria for PMDD and 22 for PMS. The groups were comparable in age, general cognitive ability, education, employment, relationship status, health habits, mental health, cycle length and scheduling of test sessions (compare **Table 1**; all p > 0.08).

**Table 1:** Demographics

|  | Control (n = 54) | PMS/PMDD (n = 51) |
| --- | --- | --- |
| Age | 24.26 ± 3.48 | 24.45 ± 3.89 |
| General Cognitive Ability [%] | 94.29 ± 7.90 | 92.97 ± 9.48 |
| Education [% Qualification for University entrance] | 96 % (n = 52) | 94% (n = 48) |
| Employment Status [% employed] | 39% (n = 21) | 36% (n = 18) |
| Relationship [% in relationship] | 43% (n = 23) | 47% (n = 24) |
| Relationship duration [years] | 3.25 ± 2.44 | 1.86 ± 2.28 |
| Smoking [% smokers] | 4% (n = 2) | 6% (n = 3) |
| BDI | 7.45 ± 5.11 | 9.51 ± 6.34 |
| BAI | 9.22 ± 5.92 | 8.25 ± 5.21 |
| Cycle duration [days] | 28.67 ± 2.54 | 28.75 ± 2.54 |
| Cycle day mid-follicular | 7.72 ± 1.13 (n = 53) | 7.04 ± 2.81 (n = 51) |
| Cycle day mid-luteal | -6.58 ± 1.92 (n = 52) | -6.78 ± 1.80 (n = 51) |
| Cycle day late luteal | -1.85 ± 0.82 (n = 46) | -1.79 ± 0.91 (n = 43) |

### Group differences and menstrual cycle related changes in hormones, mood and emotion regulation

For all group and menstrual cycle comparisons, descriptive statistics can be found in **Table 2**, statistical parameters in **Table 3**.

**Table 2:** Means ± standard deviations of hormones, mood and emotion regulation by group and cycle phase

|  | Control group |  |  | PMS/PMDD group |  |  |
| --- | --- | --- | --- | --- | --- | --- |
|  | Mid-foll. | Mid-luteal | Late-Luteal | Mid-foll. | Mid-luteal | Late-Luteal |
| Estradiol | 1.16 ± 0.60 | 1.29 ± 0.61 | 1.21 ± 0.66 | 1.18 ± 0.72 | 1.33 ± 0.44 | 1.36 ± 0.96 |
| Progesterone | 50.88 ± 52.42 | 176.32 ± 126.43 | 72.16 ± 63.22 | 53.03 ± 60.82 | 166.15 ± 104.23 | 102.94 ± 103.71 |
| DRSP Psych | 1.95 ± 0.74 | 1.73 ± 0.65 | 2.06 ± 0.71 | 1.62 ± 0.60 | 2.20 ± 0.90 | 2.73 ± 0.90 |
| DRSP Phys | 1.28 ± 0.35 | 1.35 ± 0.42 | 1.57 ± 0.53 | 1.25 ± 0.38 | 1.51 ± 0.52 | 1.84 ± 0.62 |
| DRSP Impact | 1.79 ± 0.88 | 1.47 ± 0.68 | 1.57 ± 0.68 | 1.44 ± 0.72 | 1.77 ± 1.03 | 2.26 ± 0.95 |
| Pos. Affect | 2.97 ± 0.80 | 3.24 ± 0.82 | 3.15 ± 0.68 | 3.18 ± 0.81 | 2.76 ± 0.80 | 2.43 ± 0.65 |
| Neg. Affect | 2.03 ± 0.62 | 1.89 ± 0.54 | 2.01 ± 0.59 | 1.87 ± 0.51 | 2.14 ± 0.60 | 2.43 ± 0.62 |
| STAI | 2.01 ± 0.46 | 1.94 ± 0.52 | 2.04 ± 0.44 | 1.95 ± 0.47 | 2.17 ± 0.44 | 2.22 ± 0.44 |
| PSS | 2.69 ± 0.69 | 2.56 ± 0.78 | 2.66 ± 0.64 | 2.50 ± 0.66 | 2.93 ± 0.66 | 3.18 ± 0.54 |
| ERT Reactivity | 0.92 ± 0.75 | 1.03 ± 1.04 | 0.93 ± 0.86 | 0.95 ± 0.65 | 0.88 ± 0.63 | 1.01 ± 0.73 |
| ERT Regulation | 0.22 ± 0.17 | 0.21 ± 0.17 | 0.19 ± 0.22 | 0.23 ± 0.15 | 0.16 ± 0.17 | 0.19 ± 0.17 |
| ERQ Reappraisal | 4.39 ± 1.13 | 4.61 ± 1.28 | 4.55 ± 1.07 | 4.51 ± 0.94 | 4.30 ± 1.23 | 4.14 ± 1.19 |
| ERQ Suppression | 3.18 ± 1.34 | 2.78 ± 1.13 | 3.03 ± 0.99 | 2.76 ± 1.13 | 2.95 ± 1.12 | 2.93 ± 1.20 |
| ERQ Diff | 1.21 ± 1.85 | 1.82 ± 1.92 | 1.52 ± 1.53 | 1.75 ± 1.43 | 1.35 ± 1.52 | 1.20 ± 1.66 |

**Table 3:** Statistical parameters of group and cycle comparisons

|  | Cycle Phase |  |  | Group |  |  | Cycle Phase*Group |  |  |
| --- | --- | --- | --- | --- | --- | --- | --- | --- | --- |
| | $\eta_p^2$ | F | BF<sub>01</sub> | $\eta_p^2$ | F | BF<sub>01</sub> | $\eta_p^2$ | F | BF<sub>01</sub> |
| Estradiol | 0.05 | 1.34 | 0.52 ± 1.31% | <0.01 | 0.14 | 3.63 ± 1.25% | <0.01 | 0.49 | 8.86 ± 1.73% |
| Progesterone | **0.51** | **50.33\*\*\*** | **<0.01 ± 1.20%** | <0.01 | 0.09 | 3.71 ± 1.72% | <0.01 | 0.42 | 9.45 ± 1.38% |
| DRSP Phys | **0.28** | **8.51\*** | **<0.01 ± 1.09%** | 0.02 | 0.44 | 1.65 ± 3.05% | **0.04** | **4.00\*** | 0.47 ± 2.49% |
| DRSP Psych | 0.20 | 3.10 |  | 0.03 | 5.90 |  | **0.16** | **17.64\*\*\*** | **<0.01 ± 2.25%** |
| DRSP Impact | 0.07 | 2.20 |  | 0.02 | 5.01 |  | **0.13** | **13.77\*\*\*** | **<0.01 ± 2.61%** |
| Positive Affect | 0.07 | 3.05 |  | 0.05 | 1.84 |  | **0.15** | **16.87\*\*\*** | **<0.01 ± 1.83%** |
| Negative Affect | 0.09 | 0.84 |  | 0.03 | 1.80 |  | **0.11** | **11.02\*\*\*** | **<0.01 ± 1.94%** |
| STAI | 0.05 | 0.34 |  | 0.02 | 0.35 |  | **0.04** | **4.18\*** | **0.41 ± 2.08%** |
| PSS | 0.10 | 0.60 |  | 0.04 | 1.79 |  | **0.12** | **12.60\*\*\*** | **<0.01 ± 1.46%** |
| ERT Regulation | **0.06** | **2.78** | **0.20 ± 3.54%** | <0.01 | 0.15 | 2.54 ± 4.72% | 0.03 | 3.17 | 1.31 ± 6.09% |
| ERT Reactivity | <0.01 | 0.13 | 17.32 ± 2.25% | <0.01 | 0.69 | 3.21 ± 1.97% | <0.01 | 0.57 | 9.41 ± 2.41% |
| ERQ Reappraisal | 0.02 | 0.99 | 4.85 ± 1.27% | 0.01 | 0.05 | 2.36 ± 1.83% | 0.02 | 2.24 | 2.23 ± 1.65% |
| ERQ Suppression | <0.01 | 0.78 | 24.82 ± 1.24% | <0.01 | 1.84 | 3.63 ± 1.56% | 0.03 | 1.62 | 3.60 ± 2.05% |

#### Hormone levels

As expected, both frequentist and Bayesian analyses of interaction effects confirm that there were no group differences in the trajectory of hormone levels along the menstrual cycle. Progesterone levels peaked during the mid-luteal cycle phase (mid-lut. vs. mid-foll: d = 1.21, z = 9.98, p < 0.001; mid-lut vs. late-lut: d = -0.77, z = -5.95, p < 0.001), while estradiol levels only showed a marginal increase (**Figure 1**).

**Figure 1:**
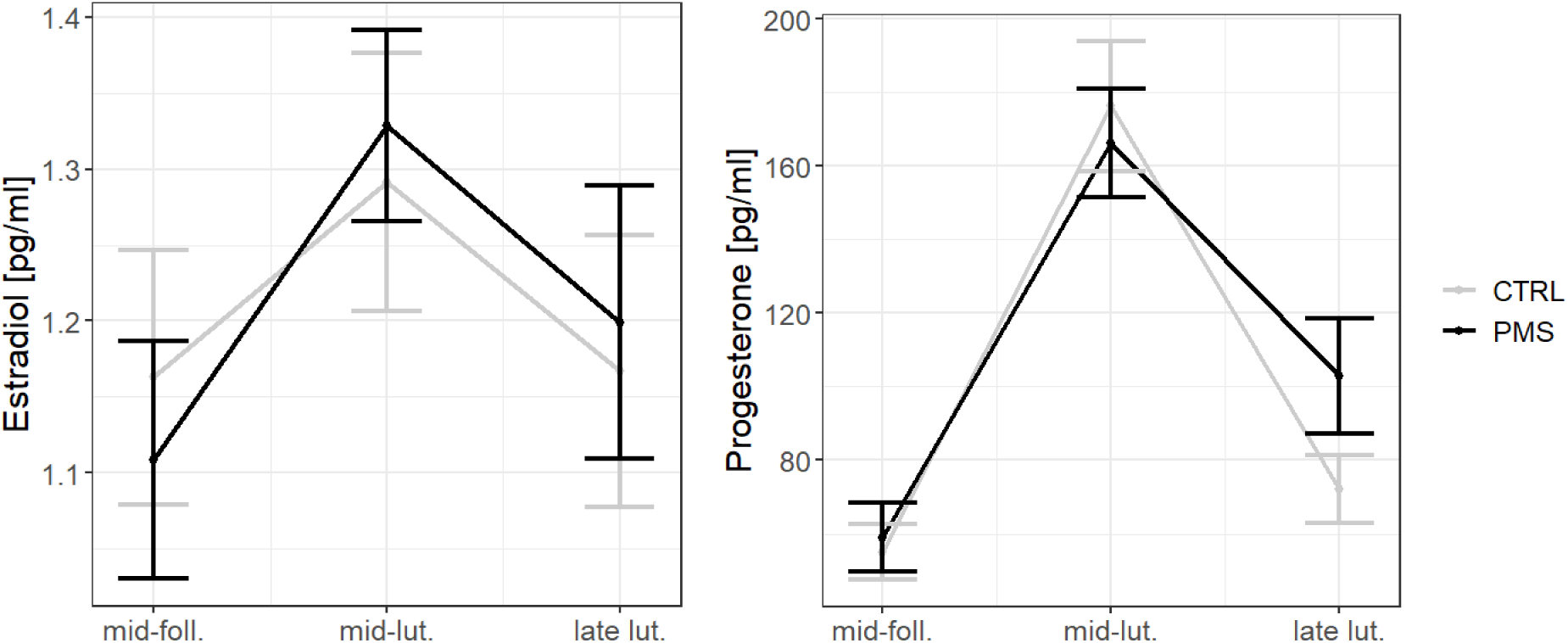
Estradiol and progesterone trajectories along the menstrual cycle. Error bars represent standard errors. Mid-foll. = mid-follicular phase, mid-lut. = mid-luteal phase, late-lut. = late-luteal phase, CTRL = control group, PMS = PMS/PMDD group.

#### Premenstrual symptoms

For *physical symptoms* there was a significant main effect of cycle phase, which interacted only weakly with groups. Physical symptoms increased significantly from the mid-follicular to late-luteal phase of the menstrual cycle in both groups (control: d = 0.71, z = 4.40, p < 0.001; PMS/PMDD: d = 1.12, z = 6.70, p < 0.001), though only the PMS/PMDD-group already experienced symptoms in the mid-luteal phase (control: d = 0.20, z = 1.29, p = 0.40; PMS/PMDD: d = 0.50, z = 3.11, p = 0.005; compare Figure 2A). Symptoms did not differ between groups in the mid-follicular phase (d = 0.10, t_(94)_ = 0.51, p = 0.61). For *psychological symptoms*, there were no significant main effects of group or menstrual cycle, but a significant interaction between group and menstrual cycle. In the PMS/PMDD group, psychological symptoms increased significantly from mid-follicular to the mid-luteal phase of the menstrual cycle (d = 0.57, z = 3.65, p < 0.001), as well as from the mid-luteal to the late-luteal phase (d = 0.64, z = 3.93, p < 0.001; compare Figure 2B). In the control group, there were no significant changes from the mid-follicular to the mid- or late luteal phase (both |d| < 0.27, both |z| < 1.77, both p > 0.18), though Bayesian evidence was only anecdotal in that respect (BF_01_ = 1.95 ± 0.92%). Interestingly, women with PMS/PMDD reported less psychological symptoms than women in the control group during the mid-follicular phase (d = 0.49, t_(96)_ = 2.45, p = 0.02).

**Figure 2:**
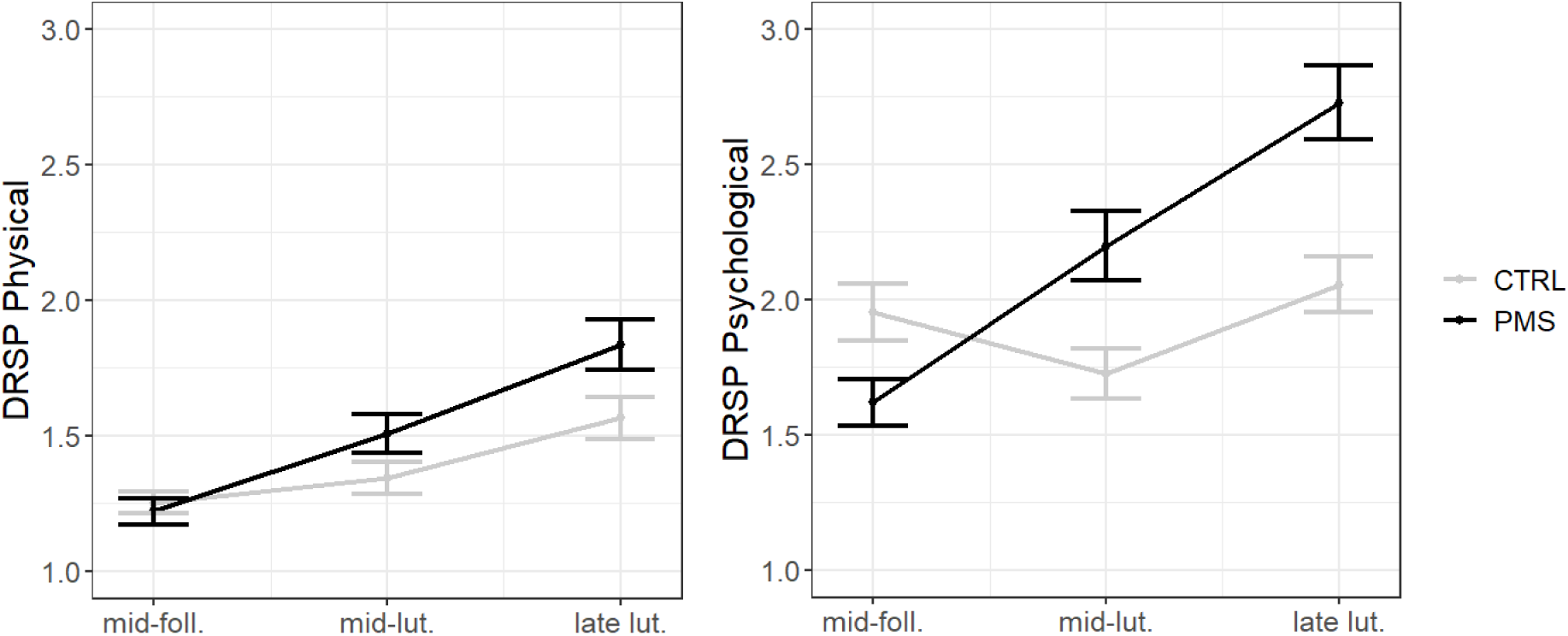
Premenstrual symptom trajectories along the menstrual cycle. Error bars represent standard errors. Mid-foll. = mid-follicular phase, mid-lut. = mid-luteal phase, late-lut. = late-luteal phase, CTRL = control group, PMS = PMS/PMDD group.

Impact of premenstrual symptoms on everyday life followed a similar pattern as psychological symptoms, though changes in impact in the PMS/PMDD group emerged only after the mid-luteal phase (mid-lut. vs. mid-foll.: d = 0.31, z = 1.73, p = 0.19; late-lut. vs. mid-lut.: d = 0.55, z = 2.94, p = 0.009) with no changes in the control group (all |d| < 0.34, all |z| < 2.14, all p > 0.08; BF_01_ = 1.53 ± 0.99%). Mediation analyses confirmed that adding physical symptoms to the linear mixed model evaluating impact on daily life, the interaction between cycle phase and group remained significant (F_(2, 178)_ = 10.44, p < 0.001). However, after adding psychological symptoms, the interaction effect between cycle phase and group disappeared (F_(2, 178)_ = 2.34, p < 0.10), suggesting that psychological, but not physical symptoms explained group and menstrual cycle differences in impact on daily life.

#### Mood

Premenstrual symptom patterns were reflected in all mood measures with no main effects of cycle phase or group, but significant interactions of group by cycle phase (Figure 3). The PMS/PMDD group displayed a significant decrease in *positive affect* from the mid-follicular to the mid-luteal phase (d = -0.47, t = -3.20, p = 0.004) and from the mid-luteal to the late-luteal phase (d = -0.45, z = -2.85, p < 0.02), while the control group even showed an increase in positive affect from the mid-follicular to the mid-luteal phase (d = 0.36, z = 2.5, p = 0.03), with no changes from the mid-luteal to the late-luteal phase (d = -0.15, z = -1.03, p = 0.55). There were no significant group differences in positive affect in the mid-follicular phase (d = -0.26, t_(102)_ = -1.35, p = 0.18). Vice versa, in the PMS/PMDD group *negative affect*, *state anxiety (STAI)* and *perceived stress (PSS)* increased significantly from the mid-follicular to the mid-luteal phase (negative affect: d = 0.43, z = 2.50, p = 0.03; STAI: d = 0.39, z = 2.33, p = 0.05; PSS: d = 0.60, z = 3.71, p < 0.001) and negative affect and perceived stress also from the mid-luteal to the late-luteal phase (negative affect: d = 0.50, z = 2.73, p = 0.02; STAI: d = 0.25, z = 1.38, p = 0.35; PSS: d = 0.40, z = 2.32, p = 0.05), with no changes along the menstrual cycle in the control group (all |d| < 0.16, all |z| < 1.24, all p > 0.43; all BF_01_ > 7.57). There were no significant group differences in negative affect, state anxiety or perceived stress in the mid-follicular phase (all |d| < 0.30, all |t| < 1.51, all p > 0.13).

**Figure 3:**
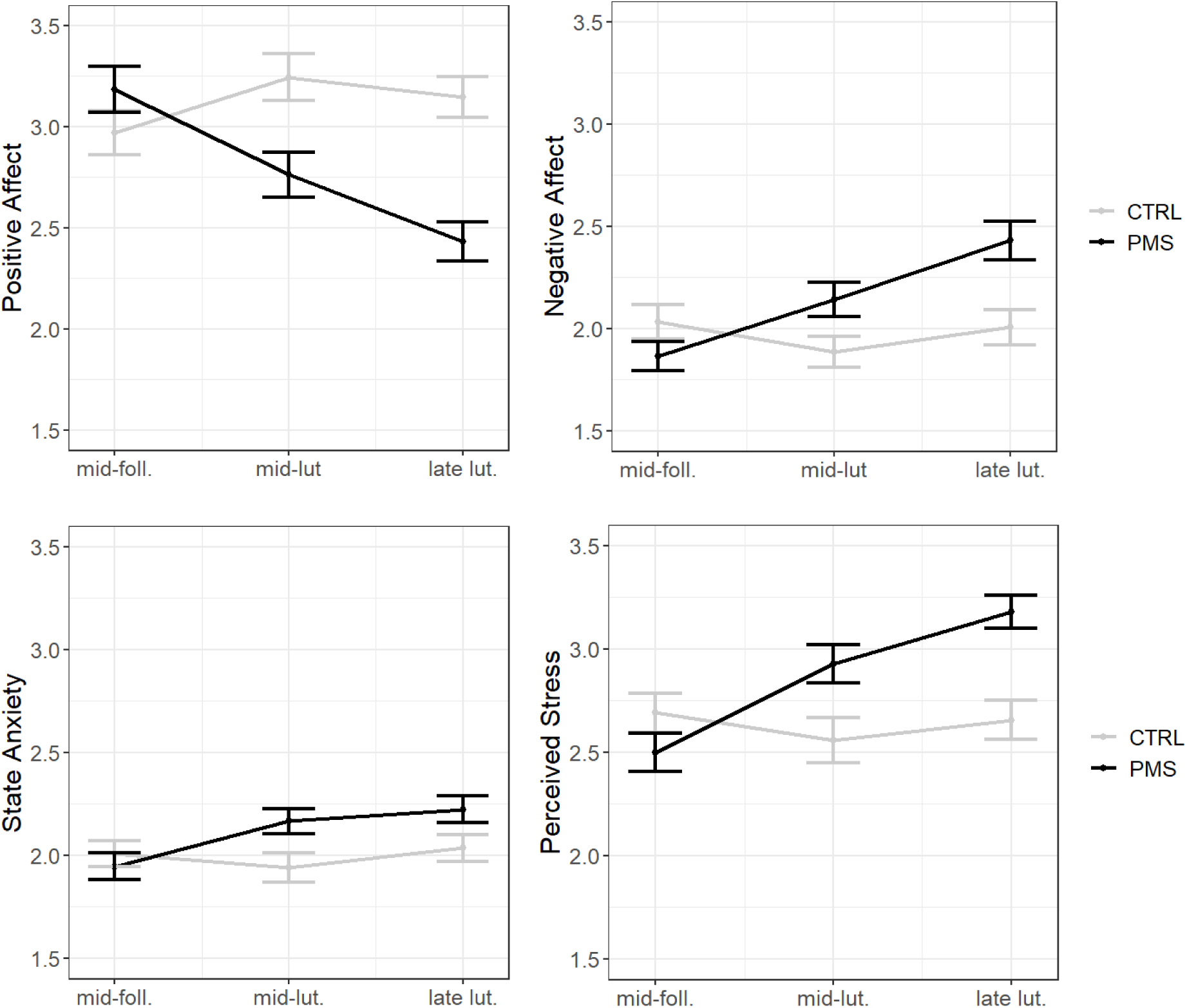
Mood trajectories along the menstrual cycle in women with and without PMS/PMDD. Error bars represent standard errors. Mid-foll. = mid-follicular phase, mid-lut. = mid-luteal phase, late-lut. = late-luteal phase, CTRL = control group, PMS = PMS/PMDD group.

#### Emotion regulation

*Difficulties in emotion regulation (DERS)* were significantly increased (d = -0.37, t = -2.34, p = 0.023; Figure 4A) in the PMS/PMDD group (M = 2.49, SD = 0.60) compared to the control group (M = 2.27, SD = 0.60) and significantly related to psychological symptom strength (r = 0.47, t = 4.00, p < 0.001; Figure 4B).

**Figure 4:**
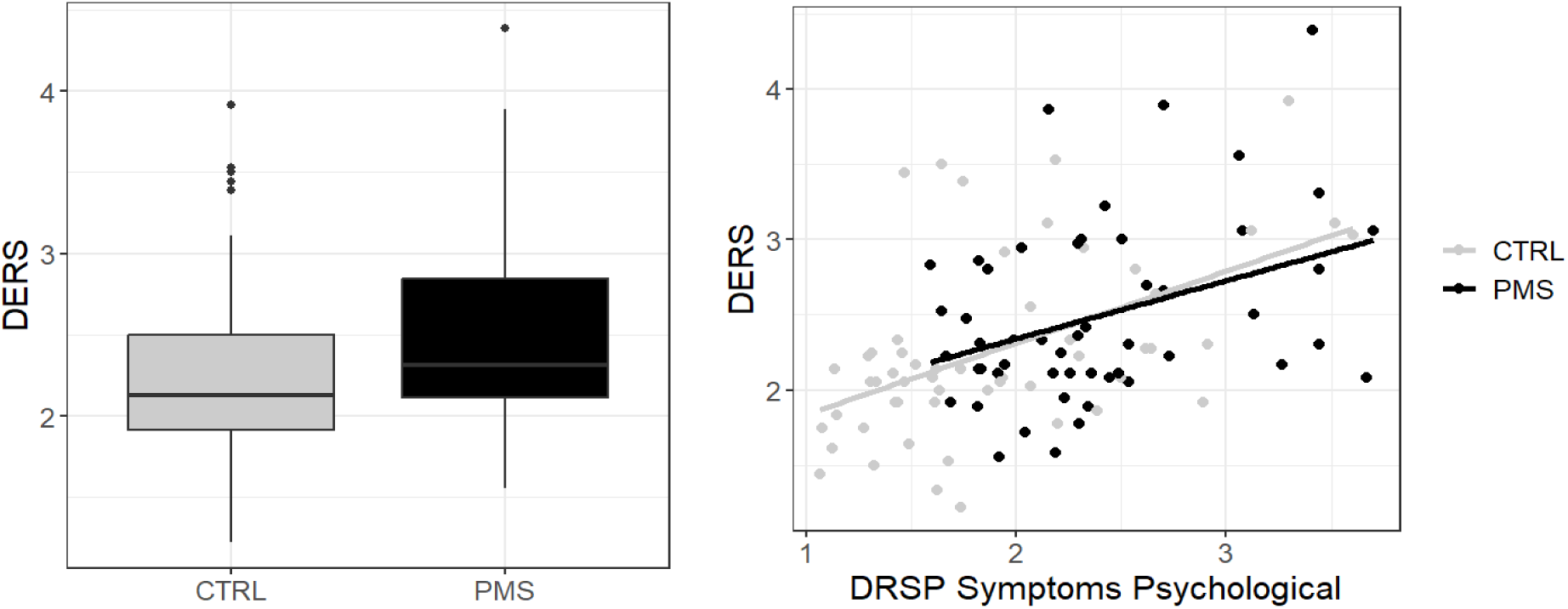
**Association of premenstrual symptoms with difficulties in emotion regulation (DERS).**

In the *emotion regulation task*, there were no group differences and no significant interaction between cycle phase and group in either emotion regulation or emotional reactivity (compare **Table 3**). Bayesian analyses suggest that a model including cycle phase is five times more likely than a model without cycle phase, though the main effect of cycle phase emerged only as trend in the frequentist statistics, with reduced emotion regulation efficacy in the late luteal compared to the mid-luteal cycle phase (d = - 0.26, z = -2.31, p = 0.05). In the *emotion regulation questionnaire,* neither reappraisal nor suppression were modulated by group, menstrual cycle or their interaction (compare **Table 3**).

**Table 3:**
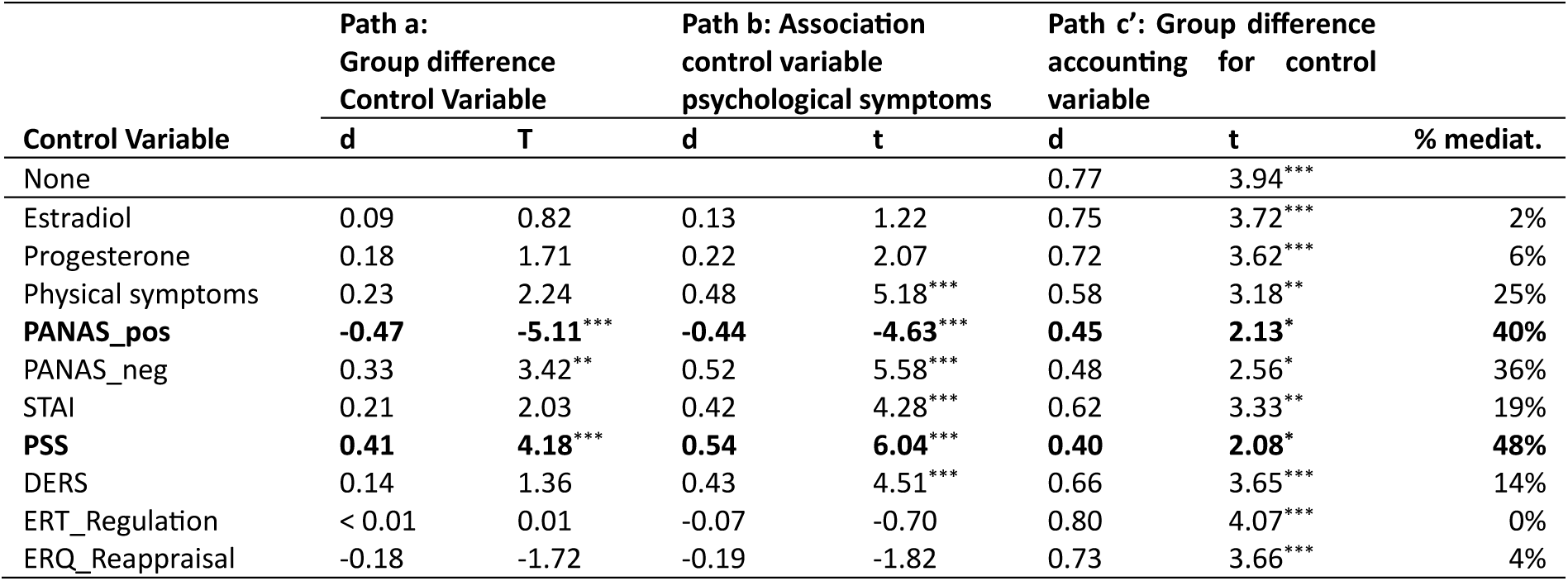
Mediation of premenstrual symptoms

### Explaining psychological symptoms

To evaluate the overlap between psychological symptoms in the late luteal phase and other variables, mediation analyses were conducted. **Table 4** lists the results of mediation analyses effect sizes. No variable could fully account for group differences in psychological symptoms. Specifically, neither hormones, nor physical symptoms nor emotion regulation variables explained the group differences in psychological symptoms. However, a partial overlap was observed with other mood variables. Specifically, the highest proportion mediated was observed for positive affect and perceived stress.

## Discussion

The current study was designed to address whether women with PMS/PMDD displayed problems with emotion regulation in comparison to healthy controls and whether these problems were trait-like (i.e., irrespective of cycle phase) or restricted to a specific cycle phase. In case of the latter, we were specifically interested, whether the problems were restricted to the symptomatic premenstrual phase or whether the problems already emerged during the mid-luteal cycle phase, when progesterone levels peak. While we were able to confirm subjectively reported difficulties in emotion regulation in women with PMS/PMDD on an extensive trait questionnaire, none of the state measure employed in the current study yielded differences in emotion regulation between women with PMS/PMDD and healthy controls in any cycle phase. However, the most important findings of the current study lie in the pattern of premenstrual symptoms. In the following sections, we will first discuss the results regarding our research questions on emotion regulation and subsequently explore the pattern of mood changes along the menstrual cycle in relation to PMS/PMDD.

Our findings regarding the difficulties in emotion regulation scale (DERS) were in accordance with previous studies (Petersen et al., 2016; Manikandaman et al., 2016; Reuveni et al., 2016; Elazar et al., 2023). Women with PMS/PMDD reported more difficulties in emotion regulation. Notably, we were even able to demonstrate that the extent of the reported difficulties increased with the severity of premenstrual symptoms. Exploratory analyses confirmed this association across all DERS subscale scores (all r > 0.31, all t > 3.11, all p_FDR_ < 0.001), including emotional clarity, emotional awareness, goal-directed behavior, impulse control, emotion regulation strategies, and non-acceptance of emotional reactions (Gutzweiler & In-Albon, 2019). This lack of domain specificity suggests that future research might benefit from utilizing a short form of the DERS for situational assessment of emotion regulation difficulties in women with PMS/PMDD.

Importantly, even though women subjectively reported more difficulties in emotion regulation, irrespective of cycle phase compared to controls, these difficulties were not reflected in emotion regulation efficacy in an experimental task (ERT) or to differences in state reports regarding the situational use of reappraisal or suppression, as measured by the emotion regulation questionnaire (ERQ). While the absence of differences in the ERQ could be attributed to the DERS’s broader scope, as it covers more emotion regulation strategies than just reappraisal or suppression, this argument does not hold for the ERT, where participants received no specific instructions on which emotion regulation strategy to employ. With the exception of Petersen et al. (2016) who found no differences between women with PMDD and healthy controls in the ERQ, previous studies employing the ERQ have generally shown reduced reappraisal in women with PMS/PMDD (Wu et al., 2016; Nayman et al., 2022; Nasiri et al., 2022; Lin et al., 2022). However, these studies used the ERQ as a trait measure, assessing habitual use of reappraisal and suppression, whereas our study utilized it as a state measure and focused on situational use. As a trait measure, the ERQ bares similarities to the DERS and thus yields similar results. Consistent with our findings, other studies have also failed to find group differences in subjective reports of situational reappraisal (Wu et al., 2016).

There are two potential explanations for these discrepancies between state and trait measures of emotion regulation. On the one hand, it is possible that women’s subjective accounts do not accurately reflect the emotion regulation ability they are in fact able to display in a given situation. Women with PMS/PMDD might perceive the severity of their premenstrual symptoms as a failure to regulate their own emotions, when it is simply a heightened challenge to their emotion regulation abilities. In line with this hypothesis, difficulties in emotion regulation only accounted for 14% of the group differences in premenstrual symptoms in the late luteal cycle phase, raising questions about the directionality of the association between premenstrual symptoms and subjective emotion regulation ability. It remains unclear, whether difficulties in emotion regulation underly premenstrual symptoms or arise as a consequence of a lifetime of untreated PMS/PMDD.

On the other hand, the emotion regulation task employed in the current study may not have been challenging enough to pick up on group differences in emotion regulation ability, while emotionally challenging situations in everyday life may be more demanding. Obviously, participants’ reports of emotional reactivity to emotional images were still subjective and as such could not be objectively confirmed, which is an inherent problem to behavioral emotion regulation tasks. However, despite these design limitations, the task was sensitive enough to detect small changes in emotion regulation performance related to the menstrual cycle in the Bayesian analysis approach. Irrespective of the intensity of premenstrual symptoms, emotion regulation efficacy was impaired during the premenstrual phase. While previous studies gave some indication that subjective reappraisal efficacy drops across the menstrual cycle (Wu et al., 2014; Doornweerd et al., 2024), these studies did not include the premenstrual phase. This may also explain, why previous studies did not observe menstrual cycle changes in the emotion regulation task (Doornweerd et al., 2024). However, taken together the evidence of the current study suggests that if there are objective differences in emotion regulation ability between women with PMS/PMDD and healthy controls, they must be subtle and probably require an emotionally highly demanding paradigm to be detectable at the behavioral level. Thus, in order to determine whether emotion regulation impairment in PMS/PMDD is a trait or emerges as state during specific cycle phases, it may be more fruitful to pursue the topic via psychobiological markers of emotion regulation effort, as has been attempted in Peterson et al. (2016).

Turning to the pattern of premenstrual symptom changes along the menstrual cycle the most noteworthy result of the current study is that unlike physical symptoms, psychological symptoms and mood worsening during the premenstrual phase are not a universal occurrence. Mood worsening was not only more severe in the PMS/PMDD group, but was in fact absent in the control group as confirmed by Bayesian analyses. While group differences in the severity of symptoms were expected per study design, the fact that no symptoms were observed in the control group warrants particular attention. Several investigations have reported mood changes along the menstrual cycle without differentiating between women with and without PMS/PMDD (Pletzer et al., 2024; Pierson et al., 2021). These results are often interpreted as seemingly demonstrating that mood changes in the premenstrual phase are a normal occurrence for women irrespective of their PMS/PMDD status. The results of our study, as well as previous investigations (Petersen et al., 2016) challenge this interpretation and suggest that these previous results may have been driven by the proportion of women in the samples suffering from PMS/PMDD, while the majority of participants, i.e., an estimated 75% based on prevalence rates (Halbreich et al., 2003), were unaffected. In light of this finding, the fact that in Western countries women rarely receive a diagnosis of PMS/PMDD by a clinician (Weisz et al., 2021), appears to be particularly problematic. In the current study, only two women of the symptomatic group had ever received a diagnosis of PMS/PMDD by a clinician, highlighting a lack of clinical attention to this syndrome. The assumption that mood changes along the menstrual cycle are common, represents a dangerous double-bind for women. While asymptomatic women may be stereotyped and experience discrimination due to assumed mood changes, many symptomatic women never receive a diagnosis and accordingly also don’t receive treatment for their symptoms, even though they severely impact their quality of life.

The validity of the PMDD diagnosis is further supported by the finding that psychological symptoms are not merely an emotional reaction to physical symptoms. Unlike psychological symptoms, premenstrual physical symptoms were reported by both groups. However, even though physical symptoms were slightly elevated in women with PMS/PMDD, the differences to the control group were not significant and physical symptoms only accounted for 25% of the psychological symptoms in a mediation analysis.

However, mediation analyses did confirm that premenstrual symptoms strongly overlap with positive affect and perceived stress, which explains why group differences were strongly reflected in these mood measures.

Finally, it is noteworthy, that premenstrual symptoms and mood worsening already emerged during the mid-luteal cycle phase, which is in line with progesterone triggering these emotional changes (Sundstrom-Poromaa et al., 2020). However, women with PMS/PMDD also appear to profit from a reduction in premenstrual symptoms during the mid-follicular phase, when their symptom burden is in fact lower than in the control group. It is possible that this finding reflects the beginning of a mood heightening during the peri-ovulatory phase. It can thus be speculated, that women with PMS/PMDD may not only be particularly vulnerable to mood-worsening throughout the luteal phase but may also profit more than control women from a mood heightening during the peri-ovulatory phase, which also heightens the contrast between positive and negative mood states across cycle phases. If this is in fact the case, the role of estradiol in triggering these mood improvements and preparing progesterone actions should be further explored.

In summary, the current study demonstrates that women subjectively experience difficulties in emotion regulation associated with premenstrual symptoms, but objective impairments in emotion regulation efficacy were not detectable with the paradigm employed in the current study. Mood changes were already observed during the mid-luteal cycle phase and absent in the control group, highlighting the importance for accurate diagnosis and treatment of PMS and PMDD.

## Data Availability

All data produced in the present study are available upon reasonable request to the authors.

## Acknowledgements

We thank all participants for their time and willingness to contribute to this study.

The work was in part funded by the European Research Council (850953) and the Austrian Science Fund (W1233).

## Notes

### Competing Interest Statement

The authors have declared no competing interest.

### Author Declarations

The ethics committee of the University of Salzburg gave ethical approval for this work (GZ 24/2022).

